# Motion Sifnos: A randomized, double-blind, placebo-controlled study demonstrating the effectiveness of tradipitant in the treatment of motion sickness

**DOI:** 10.1101/2020.04.06.20055715

**Authors:** Vasilios M. Polymeropoulos, Mark É. Czeisler, Mary M. Gibson, Austin A. Anderson, Jane Miglo, Jingyuan Wang, Changfu Xiao, Christos M. Polymeropoulos, Gunther Birznieks, Mihael H. Polymeropoulos

**Author notes:** Corresponding author (VMP). The Institute for Breathing and Sleeping, Austin Health, Heidelberg, Victoria, Australia. College of Pharmacy, University of Illinois at Chicago, Chicago, Illinois, United States of America. These authors contributed equally to this work (Mary M. Gibson, Austin A. Anderson).

## Abstract

**Background:** Novel therapies are needed for the treatment of motion sickness given the inadequate relief, and bothersome and dangerous adverse effects of currently approved therapies. Neurokinin-1 (NK1) receptor antagonists have the potential to be effective in improving the symptoms of motion sickness, given the involvement of Substance P in nauseogenic and emetic pathways and the expression of NK1 receptors in the gastrointestinal system. Here, we evaluated the efficacy of tradipitant, a novel NK1 receptor antagonist, in preventing motion sickness in variable sea conditions.

**Methods:** A total of 126 adults participated in the Motion Sifnos Study. Groups of participants were assigned to one of seven boat trips lasting approximately four hours on the Pacific Ocean. Participants were randomized 1:1 to tradipitant 170 mg or placebo and completed the Motion Sickness Severity Scale (MSSS) every 30 minutes, in addition to other assessments. Severity of motion sickness was assessed with the incidence of vomiting and the MSSS.

**Results:** Participants on tradipitant had a significantly lower incidence of vomiting as compared to those on placebo across all boat trips (tradipitant=17.5%, placebo=39.7%, p=0.0039). For trips exposed to rough sea conditions, the difference in the incidence of vomiting between the groups was more dramatic (tradipitant=15.79%, placebo=72.22%, p=0.0009). Across these trips, motion sickness symptoms were significantly lower in the tradipitant group compared to the placebo group (tradipitant=3.19, placebo=4.57, p=0.0235).

**Discussion:** Tradipitant has the potential to be an effective therapy for the prevention of vomiting and treatment of nausea in people with motion sickness.

## Introduction

Motion sickness is a disorder that has been plaguing travelers in various vehicles since antiquity [1]. Nausea and vomiting are the core symptoms of motion sickness [2]. Other symptoms may include sweating, dizziness, headache, irritability, and loss of appetite. The etiology and pathogenesis of disease are most commonly theorized as an aggregation of conflicting sensory information that leads to an unpleasant physiological reaction resulting in the symptoms described above [3-4]. Travelers with motion sickness have impaired cognitive performance [5], which can be of increased burden and danger in professional environments. Although there is a significant demand for efficacious therapies, the only currently approved treatments in the United States are dimenhydrinate (Dramamine), hyoscine (Scopolamine), and meclizine (Dramamine non-drowsy). These therapies carry incomplete efficacy and can have bothersome and dangerous adverse effects, such as drowsiness and dizziness [6]. An ideal therapy would be efficacious in the treatment of the cardinal symptoms of nausea and vomiting without inducing the array of adverse effects as seen with current medications.

NK1 receptor antagonists have the potential to be effective in the treatment of motion sickness in humans. NK1 receptors are expressed in the network of brainstem nuclei including the area postrema and nucleus tractus solitarius (NTS) that are involved in the regulation of vomiting [7]. Transmission of emetogenic sensory and emotional signals from higher cortical centers to the NTS can result in reflexive vomiting. The vestibular centers in the brainstem send signals to the NTS to trigger emesis in response to vertigo, dizziness, or visuospatial disorientation [8]. When the NTS receives the direct input from those sources, Substance P (SP), the most abundant neurokinin, is released [9]. Substance P preferentially binds to the NK1 receptors densely located in the NTS, which triggers this cascade of physiological responses, including emesis [8].

NK1 receptor antagonists are effective and approved for the treatment of nausea and vomiting in other indications. Maropitant is an approved NK1 receptor antagonist shown to be effective for the prevention of vomiting in dogs and cats due to motion sickness [10]. The NK1 receptor antagonist aprepitant is approved for the prevention of post-operative nausea and vomiting [11].

Tradipitant (VLY-686) is a novel NK1 receptor antagonist in development for the treatment of gastroparesis and atopic dermatitis. Tradipitant was shown to be effective in the treatment of nausea and vomiting in gastroparesis in a 4-week randomized study [12]. Given this evidence to support the use of tradipitant as an NK1 receptor antagonist to treat nausea and vomiting and specifically in motion sickness, we designed and conducted the Motion Sifnos Study.

Regarding the election of modality to induce motion sickness, we conducted the study in natural sea conditions in the Pacific Ocean for several reasons: (1) seasickness is the classical archetype to induce motion sickness (2) exposure to natural sea motion supports the study conditions as ecologically valid, and (3) simulation in real-world conditions offers the unique opportunity to evaluate the medication as it would be used once available to the public. Further, sea travel consists of vertical sinusoidal motions induced by waves. The amplitude of these motions is related to the wave height and corresponds to heave (linear vertical motion), which is the strongest stimulus to induce motion sickness, contributing more to provocation than the angular accelerations of pitch, roll, and yaw [13]. The frequency and acceleration of vertical sinusoidal motions can also be assessed. In an evaluation of motion sickness incidence as a function of the frequency and acceleration of vertical sinusoidal motion, wave frequencies around 0.2 Hz were most provocative, and motion sickness incidence increased with acceleration (proportional to the amplitude of the waves) across all wave frequencies [14]. The goal of the Motion Sifnos Study was to examine the effects of tradipitant in treating the symptoms of motion sickness, with a focus on the core symptoms.

## Materials and methods

The Motion Sifnos Study (NCT03772340) was a randomized, double-blind placebo-controlled study on the Pacific Ocean near Los Angeles, California to evaluate the efficacy and safety of tradipitant in the treatment of motion sickness.

### Participants

Eligible participants were adults aged between 18 and 75 years with a history of motion sickness, otherwise in good health (determined by medical and psychiatric history, physical examination, electrocardiography, serum chemistry, hematology, urinalysis, and urine toxicology) and lacking any major nausea-inducing disorders. All participants provided written informed consent. Ethical oversight of the study procedures was conducted by the Institutional Review Board at Advarra. The majority of participants had described sea travel to most severely exacerbate their symptoms of motion sickness. Recruitment of participants was accomplished through advertisements, participant databases, and pre-screening interviews conducted online or by phone script.

1270 people opted in via an online or phone Motion Sickness Eligibility Questionnaire (MSEQ) declaring interest for participation in the study. Participants eligible based on the MSEQ criteria were contacted to provide more information for the study, and interviewed about their symptoms of motion sickness to determine eligibility. A total of 126 eligible people were randomized and took part in the travel assessment. Groups ranging from 10 to 26 participants took part in a single boat trip travel assessment, among the seven that occurred in total from January to May 2019. Groups were organized based on time and scheduling availability of participants, site staff, nautical professionals, and weather conditions.

### Procedures

At Visit 1 (V1), participants were screened with medical history, physical exam, electrocardiogram, and laboratory tests. Collection of adverse event information began at the time the informed consent form was signed and continued through the end of the study. Upon arrival on the day of the boat trip (Visit 2 – V2), participants were randomly assigned (1:1 ratio) through an Interactive Web Response System and administered either two oral capsules totaling tradipitant 170 mg or two oral capsules of placebo identical in appearance approximately 60 minutes prior to initiation of the travel assessment on the boat.

Participants engaged in a travel assessment during which they spent 237 to 250 minutes on the boat in the Pacific Ocean, except where severe weather conditions limited one boat trip (Boat 5) to 148 minutes. The monohull boats were about 100 feet in length with indoor seating cabins and visibility to the outside. Each boat was temperature-controlled and chartered by a full staff of nautical professionals. Trips occurred on a different day under variable sea conditions. Duration of travel, route, average wave height, wave period, wind speed, and wind direction were recorded. The study was designed to have participants on the boat in the sea to expose them to realistic conditions to elicit symptoms of motion sickness. The stimulus was variable based on different movements induced by natural sea conditions.

On the boat, participants were instructed to remain in assigned seats, and were seated with no one siung directly adjacent to them. Participants had access to snacks including water, juice, and crackers, and could access restrooms. Every 30 minutes from the initiation of the trip participants were instructed to fill out the Motion Sickness Severity Scale (MSSS) to assess their symptoms of motion sickness. The MSSS is a 7-point scale scored 0-6 based on participant selection of symptom severity from the following choices: no symptoms, stomach awareness or discomfort, mild nausea, moderate nausea, severe nausea, retching, vomiting (Fig 1) [15]. The MSSS reflects the severity of the core symptoms of motion sickness as experienced and reported by the participant.

**Fig 1.**
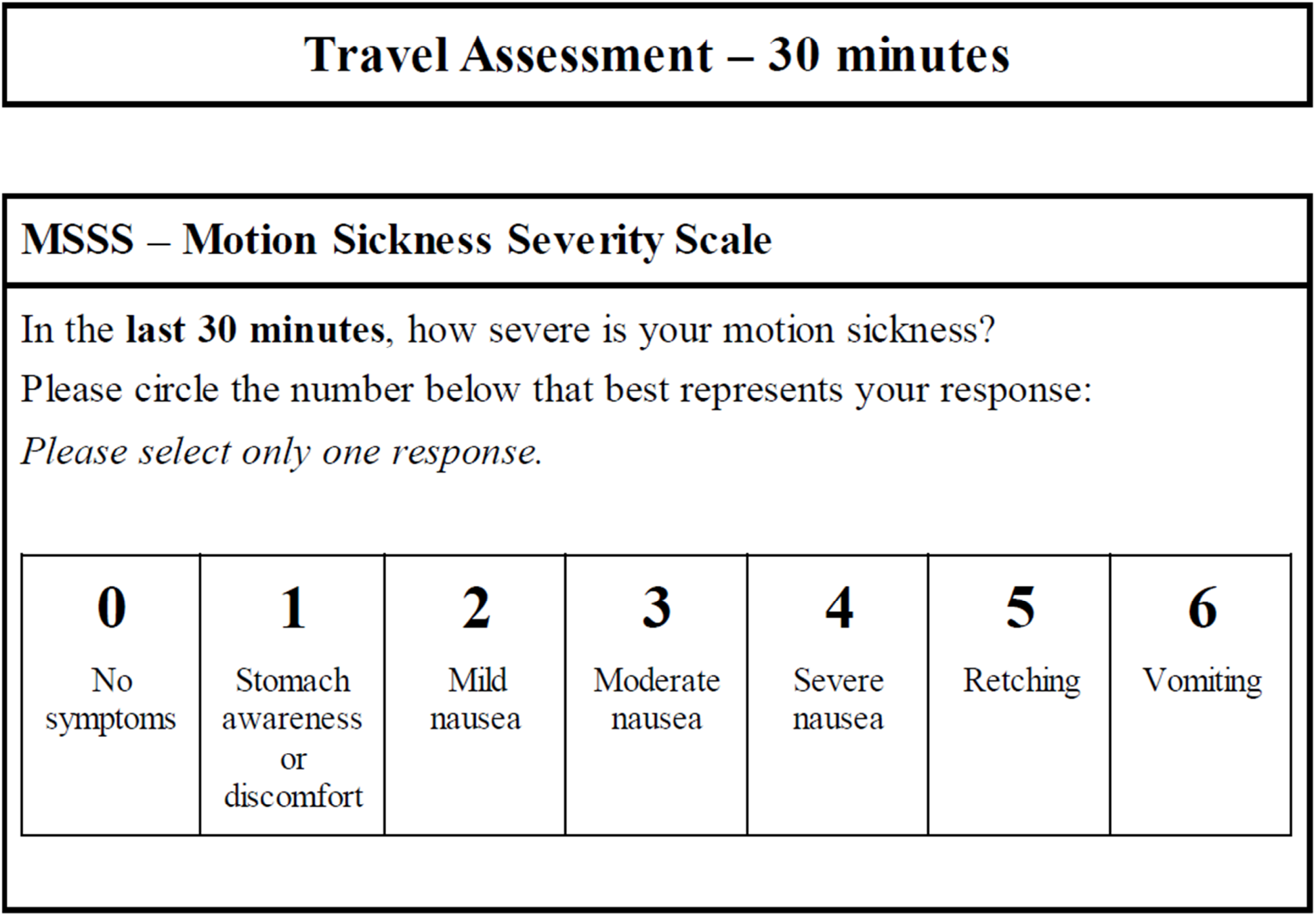
Motion Sickness Severity Scale (MSSS). The questionnaire is utilized for clarity and rapid completion (especially for participants experiencing nausea or vomiting) to facilitate repeated use over the duration of boat travel.

At the conclusion of travel, participants completed several questionnaires including the Motion Sickness Assessment Questionnaire (MSAQ) [16] and the Patient Global Impression Scale - Subjective (PGI-S) [17] for motion sickness. The MSAQ is a 16-item questionnaire ranging from 0-9 that was developed to create a validated questionnaire for motion sickness that encompasses all symptoms of motion sickness subdivided by body system, though not all of these symptoms are shared by all subjects [2,16]. The PGI-S for motion sickness is a 5-item scale rated 0-4 where subjects indicate their severity of motion sickness ranging from none to very severe.

In the event a participant vomited and felt too ill to proceed, there was an option to complete the end of study questionnaires early and move to a different area on the boat to relax and alleviate their symptoms. A synopsis of the procedure is outlined in Fig 2.

**Fig 2.**
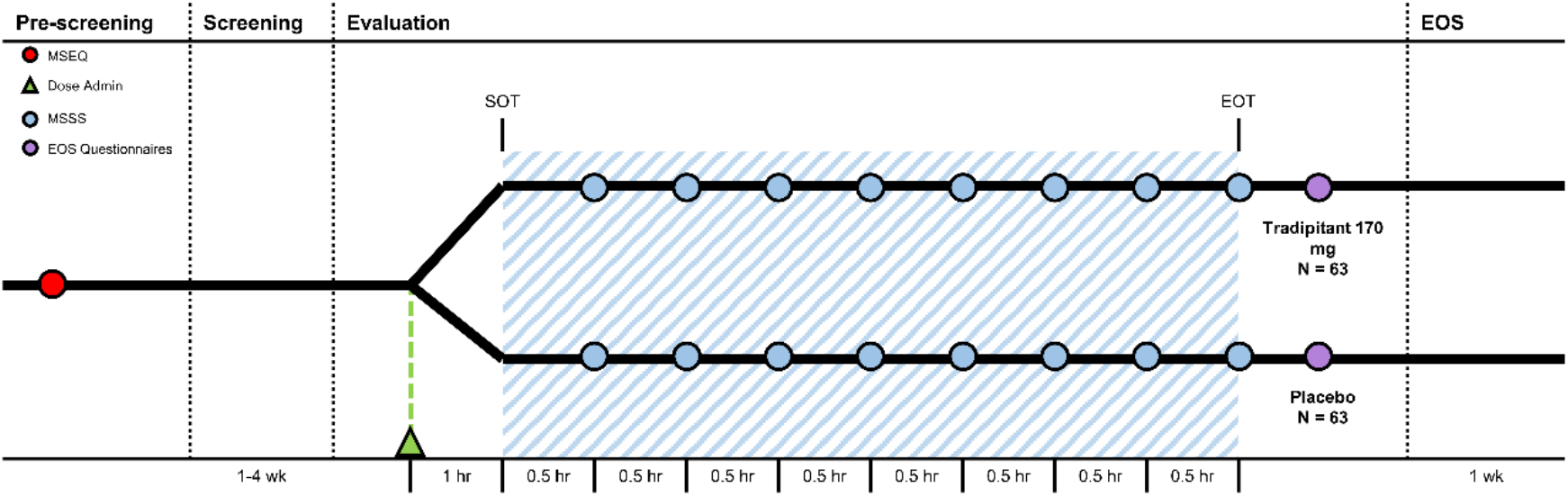
Motion Sifnos Study design. The Motion Sickness Eligibility Questionnaire (MSEQ, red circle) was used to pre-screen prospective participants for self-reported histories of motion sickness. Approximately 60 minutes prior to the start of travel (SOT), participants were randomized (Dose Administration, green triangle). The blue shaded area represents boat travel. The MSSS was administered every 30 minutes (MSSS, blue circle). Approximately 30-60 minutes aver the end of travel (EOT), participants were instructed to complete the end of study questionnaires (EOS Questionnaires, purple circle).

### Statistical analysis

There were two primary assessments for the study: the incidence (percentage) of vomiting (indicated by a score of 6 on the MSSS or by election to complete the end of study questionnaires early due to sickness severity) and the severity of motion sickness (as measured by the highest score indicated on the MSSS) during the trip. The incidence of vomiting was analyzed by a Cochran-Mantel-Haenszel (CMH) test adjusting for the trip. The most severe motion sickness severity was analyzed using an analysis of variance (ANOVA) with the main effect of treatment and boat trip. The MSAQ and PGI-S were analyzed using an ANOVA in the same manner as MSSS worst score.

Additional analyses were conducted to evaluate the treatment effect under different sea conditions given the established interaction between sea conditions and motion sickness incidence [13]. For the analyses, boat trips exposed to wave heights greater than 1.2 meters, corresponding to a Beaufort scale of 4, were considered the “Rough Sea Conditions” subgroup, and trips exposed to wave heights less than or equal to 1.2 meters were considered the “Calm Sea Conditions” subgroup.

Statistical significance was evaluated using an a priori significance level set to α=0.05. To quantify the efficacy of tradipitant in reducing vomiting incidence in rough sea conditions, the relative risk reduction (RRR) was calculated. In this context the RRR is the relative decrease in the risk of vomiting in the tradipitant group compared to the placebo group.

## Results

All randomized participants completed the study. Baseline demographic characteristics for the study population are reported in Table 1. Participants were of a diverse age, ethnicity, and physical characteristics representative of the US population. Females made up 77% of the study population, as anticipated given epidemiological data [18]. There was an average of 18 participants per boat trip. Summary statistics for the boat trip are reported in Table 2. Across trips, the frequency of vertical sinusoidal oscillation as derived from wave period was between 0.09 and 0.20 Hz. The average wave height across trips was 0.99 meters (standard deviation (sd)=0.38) and the average wind speed was 11.48 knots (sd=6.91).

**Table 1.** Baseline characteristics for the Motion Sifnos Study population.

|  | **Tradipitant 170 mg**<br>**N=63** | **Placebo**<br>**N=63** | **Total**<br>**N=126** |
| --- | --- | --- | --- |
| **Age (years)** | 38.6 (11.44) | 39.7 (12.65) | 39.1 (12.02) |
| **Sex** |  |  |  |
| *Male* | 13 (20.6%) | 16 (25.4%) | 29 (23.0%) |
| *Female* | 50 (79.4%) | 47 (74.6%) | 97 (77.0%) |
| **Race** |  |  |  |
| *American Indian or Alaskan Native* | 0 (0.0%) | 1 (1.6%) | 1 (0.8%) |
| *Asian* | 6 (9.5%) | 8 (12.7%) | 14 (11.1%) |
| *Black or African American* | 6 (9.5%) | 4 (6.3%) | 10 (7.9%) |
| *Native Hawaiian or Pacific Islander* | 2 (3.2%) | 0 (0.0%) | 2 (1.6%) |
| *White* | 44 (69.8%) | 49 (77.8%) | 93 (73.8%) |
| *Other* | 5 (7.9%) | 1 (1.6%) | 6 (4.8%) |
| **Ethnic Origin** |  |  |  |
| *Hispanic or Latino* | 22 (34.9%) | 21 (33.3%) | 43 (34.1%) |
| *Not Hispanic or Latino* | 41 (65.1%) | 42 (66.7%) | 83 (65.9%) |
| **Weight (kg)** | 74.27 (16.83) | 78.03 (16.17) | 76.15 (16.55) |
| **Body mass index (kg/m<sup>2</sup>)** | 26.58 (5.03) | 27.87 (4.86) | 27.22 (4.97) |
| **STAI-6** | 11.0 (3.84) | 11.7 (3.71) | 11.4 (3.78) |
| Baseline is defined as the last non-missing measurement prior to start of study drug. |  |  |  |
| Data are n (%), or mean (SD); STAI-6 = 6-Item Spielberger State-Trait Anxiety Inventory |  |  |  |

**Table 2.** Summary of boat trip conditions.

| **Boat Trip** | **Subjects on Boat**<br>**(n)** | **Peak Wave**<br>**Height**<br>**(m)** | **Peak**<br>**Wave**<br>**Period**<br>**(sec)** | **Average**<br>**Wind**<br>**Speed**<br>**(knots)** | **Wind**<br>**Direction**<br>**(N, S, W, E)** |
| --- | --- | --- | --- | --- | --- |
| **Boat 1: January 26, 2019** | 23 | 0.61 | 7.0 | 11.3 | WSW |
| **Boat 2: February 23, 2019** | 26 | 0.90 | 7.0 | 6.08 | SE |
| **Boat 3: March 9, 2019** | 11 | 1.22 | 4.5 | 15.0 | W |
| **Boat 4: March 30, 2019** | 16 | 1.37 | 11 | 11.0 | W |
| **Boat 5: April 9, 2019** | 10 | 1.52 | 8.0 | 25.0 | WSW |
| **Boat 6: April 30, 2019** | 18 | 0.61 | 6.0 | 7.00 | W |
| **Boat 7: May 30, 2019** | 22 | 0.67 | 5.0 | 5.00 | SW |
| Wind direction is defined as N=North, S=South, W=West, E=East |  |  |  |  |  |

A summary of assessments for the intent-to-treat (ITT) population is reported in Table 3. The incidence of vomiting in the tradipitant group was significantly lower than the placebo group across boat trips (Fig 3). In boats exposed to rough sea conditions the magnitude of difference in the incidence of vomiting was greater between groups (tradipitant=15.8%, placebo=72.2%, p=0.0009, n=37). In these boat trips with higher waves and worse sea conditions, participants taking tradipitant reduced their risk of vomiting by 78ti compared to participants taking placebo (RRR=78%, n=37).

**Table 3.**
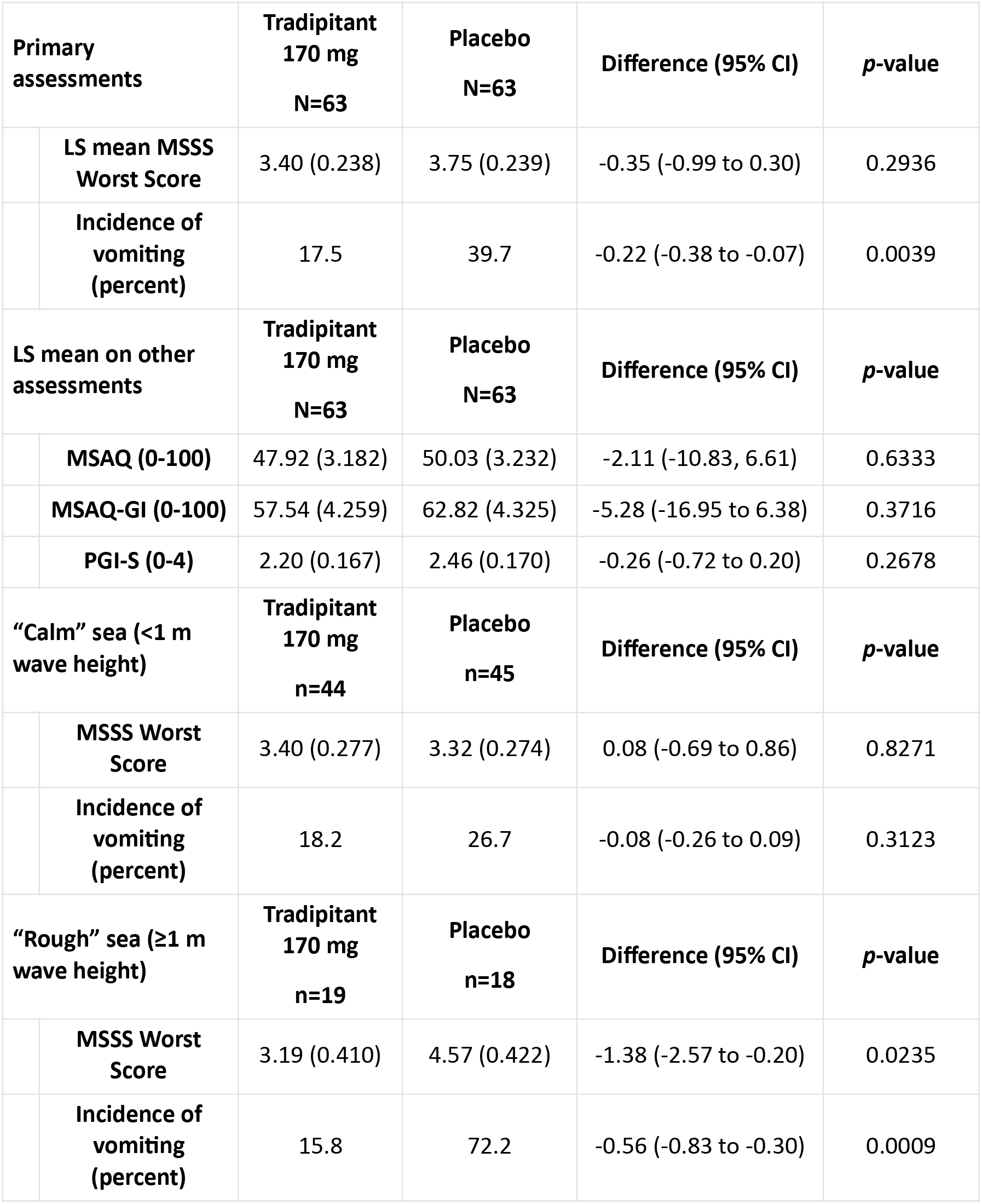
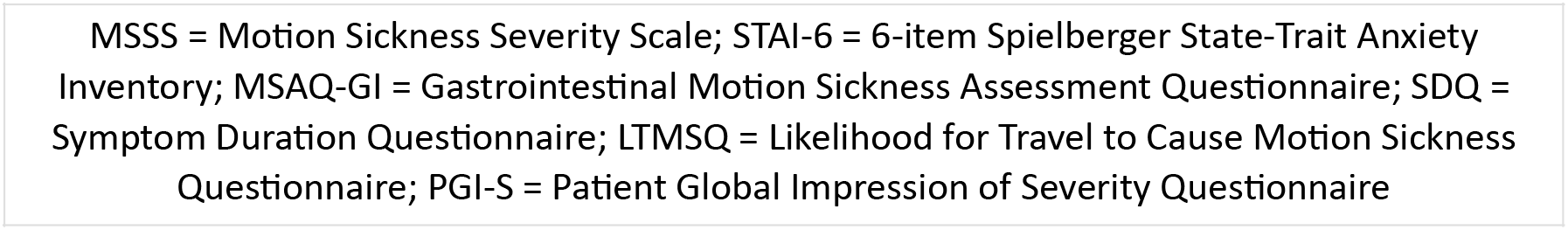
Summary of endpoints : Motion sickness symptom assessments for the ITT population.

| Primary assessments |  | Tradipitant<br>170 mg<br>N=63 | Placebo<br>N=63 | Difference (95% CI) | p-value |
| --- | --- | --- | --- | --- | --- |
|  | LS mean MSSS Worst Score | 3.40 (0.238) | 3.75 (0.239) | -0.35 (-0.99 to 0.30) | 0.2936 |
|  | Incidence of vomiting (percent) | 17.5 | 39.7 | -0.22 (-0.38 to -0.07) | 0.0039 |
| LS mean on other assessments |  | Tradipitant<br>170 mg<br>N=63 | Placebo<br>N=63 | Difference (95% CI) | p-value |
|  | MSAQ (0-100) | 47.92 (3.182) | 50.03 (3.232) | -2.11 (-10.83, 6.61) | 0.6333 |
|  | MSAQ-GI (0-100) | 57.54 (4.259) | 62.82 (4.325) | -5.28 (-16.95 to 6.38) | 0.3716 |
|  | PGI-S (0-4) | 2.20 (0.167) | 2.46 (0.170) | -0.26 (-0.72 to 0.20) | 0.2678 |
| “Calm” sea (<1 m wave height) |  | Tradipitant<br>170 mg<br>n=44 | Placebo<br>n=45 | Difference (95% CI) | p-value |
|  | MSSS Worst Score | 3.40 (0.277) | 3.32 (0.274) | 0.08 (-0.69 to 0.86) | 0.8271 |
|  | Incidence of vomiting (percent) | 18.2 | 26.7 | -0.08 (-0.26 to 0.09) | 0.3123 |
| “Rough” sea (≥1 m wave height) |  | Tradipitant<br>170 mg<br>n=19 | Placebo<br>n=18 | Difference (95% CI) | p-value |
|  | MSSS Worst Score | 3.19 (0.410) | 4.57 (0.422) | -1.38 (-2.57 to -0.20) | 0.0235 |
|  | Incidence of vomiting (percent) | 15.8 | 72.2 | -0.56 (-0.83 to -0.30) | 0.0009 |

MSSS = Motion Sickness Severity Scale; STAI-6 = 6-item Spielberger State-Trait Anxiety Inventory; MSAQ-GI = Gastrointestinal Motion Sickness Assessment Questionnaire; SDQ = Symptom Duration Questionnaire; LTMSQ = Likelihood for Travel to Cause Motion Sickness Questionnaire; PGI-S = Patient Global Impression of Severity Questionnaire

**Fig 3.**
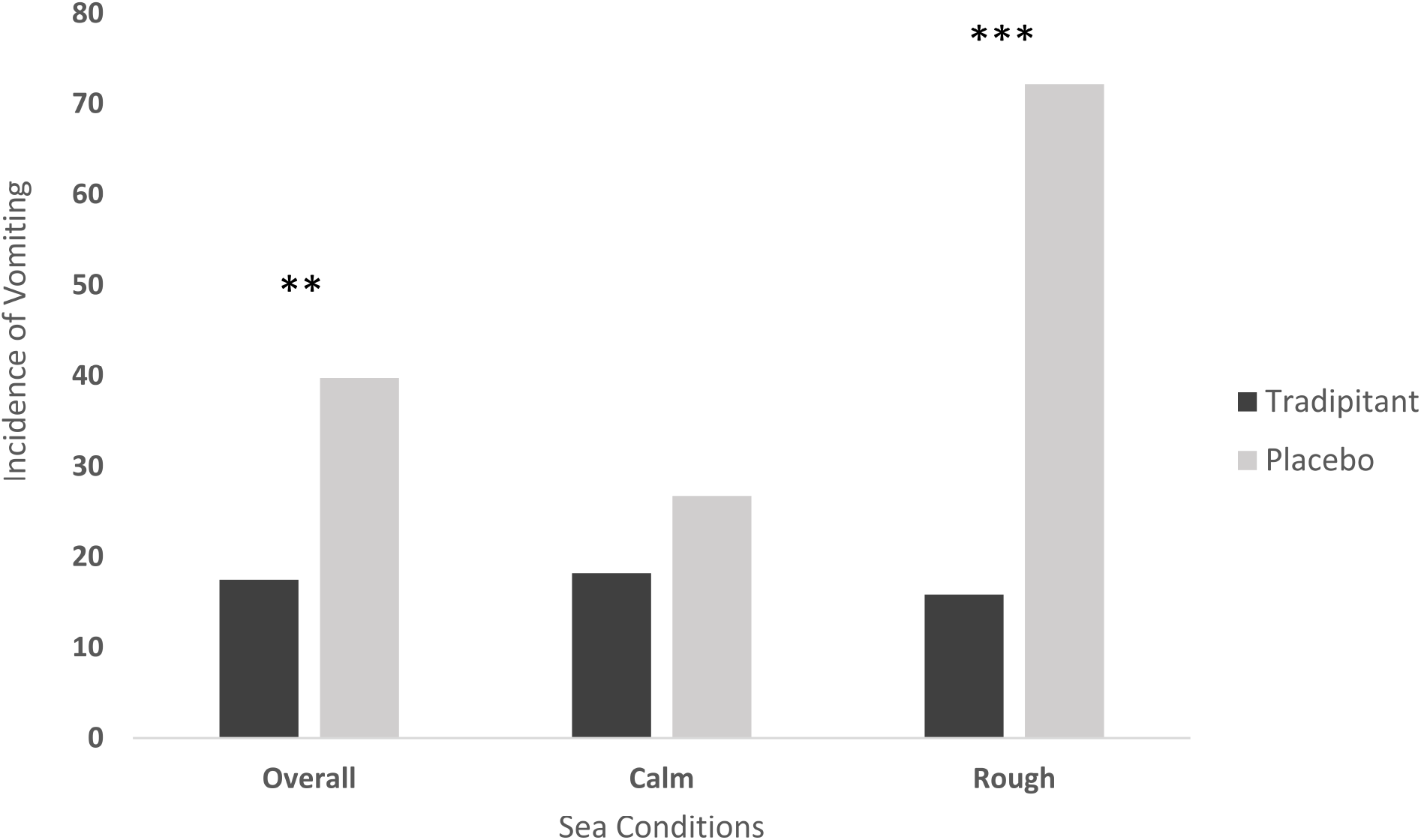
Vomiting incidence. Vomiting episodes were indicated by participants scoring a 6 on the MSSS or by election to complete the end of study questionnaires early due to sickness severity.

In a comparison of the placebo subgroups in rough versus calm sea conditions, the incidence of vomiting in participants taking placebo increased significantly in rough sea conditions (difference=45.55%, p=0.0014 [Fisher’s exact test]), as did motion sickness severity as measured by the MSSS worst score (difference=1.17, p=0.0465). In contrast, the incidence of vomiting in participants taking tradipitant remained similar in boat trips in rough and calm sea conditions (rough=15.79%, calm=18.18%, difference=2.39%, p=1.0 [Fisher’s exact test]) and the magnitude of symptoms (difference=0.32, p=0.4519) did not differ significantly.

Across all boat trips, participants on tradipitant rated their most severe motion sickness as lower compared to those on placebo (LS mean MSSS: tradipitant=3.40, placebo=3.75, p=0.2936, n=126). In rough sea conditions, tradipitant participants’ most severe motion sickness symptoms were significantly improved as compared to the placebo group (LS mean MSSS: tradipitant=3.19, placebo=4.57, p=0.0235, n=37). In calm sea conditions, participants in both groups rated their symptoms as similar (LS mean MSSS: tradipitant=3.40, placebo=3.32, p=0.8271, n=89).

In an analysis of all boat trips, assessments of MSAQ and PGI-S between treatment groups were not significantly different. When examining these assessments in boat trips in rough sea conditions, the difference between groups for the following assessments all favored the tradipitant group: MSAQ (tradipitant=49.9, placebo=59.0, p=0.2915), MSAQ-GI (tradipitant=60.3, placebo=76.4, p=0.1468), PGI-S (tradipitant=2.29, placebo=3.00, p=0.1372).

In rough sea conditions, by-item analysis of the MSAQ revealed that tradipitant was favored by greater than 1.5 points over placebo in five symptom assessments: spinning, hot/warm, sick to stomach, queasy, and faint-like.

Tradipitant 170mg was well-tolerated during the study. Tradipitant participants reported somnolence and headache more frequently than participants in the placebo group (somnolence: tradipitant=27%, placebo=12.7%, headache: tradipitant=12.7%, placebo=7.9%). No serious adverse events were reported during study participation.

## Discussion

The Motion Sifnos Study demonstrated the robust effects of tradipitant in the improvement of the core symptoms of motion sickness, particularly in rough seas. In our analyses, the consideration of the effect of variable sea conditions was important. Heave and oscillatory frequency have been established as primary stimuli of motion sickness during sea travel, with the angular acceleration of pitch, roll, and yaw contributing to a lesser degree [13]. The wave frequency of all trips was in the range of the most provocative vertical sinusoidal oscillatatory frequencies, which is typically around 0.2 Hz [14]. Although heave motion and acceleration of vertical sinusoidal oscillation were not directly recorded, increased wave height (which likely correlates with increased vertical displacement) was associated with increased motion sickness severity. Participants in the placebo group experienced a significantly higher incidence of vomiting and worsening of symptoms under these highly provocative conditions with increasing wave heights, consistent with the literature [19]. Notably, the incidence of vomiting and magnitude of symptoms remained similarly low in tradipitant participants across sea conditions.

In this study, by studying various sea conditions, we also demonstrate and validate a model for studying motion sickness effectively using wave height as a core component of the model. The findings suggest that rough sea conditions as indicated by wave height have a direct effect on the utility of sea travel as a model for studying motion sickness.

Overall, limited adverse effects were reported. Participants on tradipitant reported a low rate of adverse effects, generally reported as mild in nature. This stands in contrast to other approved therapeutics that carry significant and hazardous adverse effects.

The presumed mechanism of tradipitant would be to treat the core symptoms of motion sickness by acting at the level of the NK1 receptors in the brainstem to prevent vomiting and at the NK1 receptors in the gut influencing nauseogenic pathways and gastric motility. Mitigation of these symptoms as assessed by the MSSS supports the efficacy of tradiptiant in improving these symptoms. Additionally, though not significant, there was a greater than 1.5-point improvement on several specific symptoms in the MSAQ in addition to core symptoms, indicating this therapeutic may employ alternative treatment pathways for motion sickness by affecting vestibular processing.

Tradipitant if available could be of immediate utility to civilian travelers and for both professional and military use. This study demonstrated the potent anti-emetic effect of tradipitant that may be of significant benefit to the general public and in at-risk occupational seungs including astronauts who suffer from severe motion sickness in the first few days of travel, especially given that vomiting in the space suit can pose a significant hazard [20]. In other professional environments including on sea and in the air, tradipitant may be a useful primary or adjunct therapy given its demonstrated efficacy and mild adverse effect profile compared to currently available pharmacological therapies.

## Data Availability

All datasets included in this study are in the article/supplemental files.

## Acknowledgements

We thank the participants of the study for their commitment to helping us in find a new treatment for motion sickness. We thank Dr. Daniel Norman MD and the site study staff. We thank the nautical staff involved in the boat travel assessment. We thank Dr. Behrang Keshavarz PhD for his guidance. We thank our colleagues at Vanda Pharmaceuticals for their support in conducting the study including: Suzanne Quebbeman, Xian Cui, Patrick Collins, Pan Wang, Grace Li, Zibo Wang, and Katerina Polymeropoulos. This work was sponsored by Vanda Pharmaceuticals.

